# A statistical forecast of LOW mortality and morbidity due to COVID-19, in ARGENTINA and other Southern Hemisphere countries

**DOI:** 10.1101/2020.04.20.20072488

**Authors:** Cesar A. Barbero

**Affiliations:** Departamento de Química, Facultad de Ciencias Exactas, Fisicoquímicas y Naturales, Universidad Nacional de Rio Cuarto; Instituto de Investigaciones en Tecnologías Energéticas y Materiales Avanzados

## Abstract

A set of open source programs in Python is devised to fit a parametric integrated Gaussian equation to cumulative deaths due to COVID-19 in Southern Hemisphere countries. The programs were successfully tested using data from advanced outbreak trajectories (Italy and Spain). The procedure was applied to data reported by **Argentina. The projected total death toll will be 182 (277-182) with a peak of deaths (6(+/-2)) the 14 of April. The outbreak begins the 9**^**th**^ **of March and end completely the 20**^**th**^ **of May. However, already on 1**^**st**^ **of May, 2 s (95**.**45%) of the deaths have occurred. The death toll arises from a number of infected individuals between 36412 and 2275**. Then, they were to use to process data from several Southern Hemisphere countries: Argentina, Brazil, Mexico, Peru, Colombia, Ecuador, Cuba, Chile, Panama, Australia, Bolivia, Honduras, New Zealand, Paraguay, Guatemala, Venezuela, Uruguay, El Salvador, Jamaica, Haiti, Costa Rica and Nicaragua. The trend is to show low number of total deaths compared with other disease outbreaks. A total projected number of deaths between 15148 and 9939 deaths for a total population of ca. 664 M inhabitants. The projected death toll is much lower (5-10 times) than those forecasted by the Imperial College Group (ICG) even considering the best scenario of total suppression of virus transmission. Using actual mortality rates it is possible to back calculate which number of infected individuals would produce such mortality. The calculated number of infected individuals (worst case scenario) is below 2.5 million. This is significantly lower than that calculated by ICG (> 45 millions). In most countries the outbreak will end in May or early June. The dynamics of the outbreaks seems to do not saturate the health services (hospital beds) but only Peru, Ecuador and Panama should have not enough ICU beds for grave COVID-19 patients.

## Introduction

The COVID-19 disease, caused by the SARS-Cov-2 virus, is presently causing the most important crisis of world society and governance in the XXI century. The International Monetary Fund predicts a decrease of world’s product in the order of 3% in 2020,[1] While this is a dire forecast for all countries it is especially difficult for developing countries like most of those of the Southern Hemisphere, where the economy weak before the outbreak.

The World Health Organization declares COVID-19 a pandemic 13 of March.[2] However, at present number of deaths is of ca. 150.000[3], while grave or critical cases are in the order of 60.000 [3]. The number of reported cases is in the order of 2,2 million.[3] Those numbers represent a small amount (0,03%) of the world’s population and are smaller than other viral pandemics/endemics. It is estimated that up to 44.0 million people globally were living with HIV in 2018, with up to 1,1 million deaths.[4] Seasonal influenza caused 5 million cases worldwide, with 650000 deaths. However, COVID-19 pandemic is at early stages and the number of cases and deaths could grow considerably. Therefore, the capability of forecasting relevant parameters (death, critical or grave cases) is critical.

At the same time, the global response to this pandemic has been swift and harsh with shutdown of all international travel and several countries in different degrees of lockdown. The socioeconomic consequences of such response are catastrophic, such as high unemployment, businesses bankruptcies, etc. Obviously is the response, based mainly on non-pharmacological interventions(NPI),[5] the cause of the economic crisis, not the disease itself. It is reasonable to ponder why such harsh measures has been taken in response to COVID-19 and not before upon seasonal influenza, HIV, H1N1 influenza,[6] etc. The usual answer is that COVID-19 is radically different and requires such measures. Besides, there is no vaccine for COVID-19 available. However, specifically in the case of influenza H1N1, the vaccine was produced only 5-6 months after the declaration of pandemic (June 2009) and 7 months after the outbreak (April 2009).[7] In the meantime, especially in the southern hemisphere, which was in autumn-winter where influenza strains are specially active, some kind of social distancing and some antivirals were the only way to fight the disease.[8] Before the H1N1 pandemic, declared by WHO in June 2009,[9] WHO predicted 2–7 million deaths in the ‘best case’ scenario.[10]. Other expert predictions, based on the death toll pandemic influenza in 1918-1920 range from 62 million deaths,[11] to 180–360 million deaths.[12] The final death toll was below 20.000.[9] The point is relevant because the same analogies are used today to predict the outcome of COVID-19 pandemic.

In the best of my knowledge, such dire predictions do not motivate the governments at the time to enact full lockdown or even complete international travel shutdown. There were targeted flight bans and in some countries school shutdown or phase out to springtime. Obviously, the social and economic consequences were small. It is interesting that the CDC guidelines,[13] suggest social distancing measures (school closing, banning public gatherings, isolation of infected, etc.) but no lockdown. The rationale behind social isolation is grounded on the studies of the effect of such interventions on the mortality of the 1918-1919 influenza pandemic.[14,15] The community NPIs recommended by the CDC,[13] involve: School Closures and Dismissals and Social Distancing Measures for Schools, Workplaces, and Mass Gatherings. Those are made with together with personal NPI: voluntary home isolation (i.e., staying home when ill or self-isolation, respiratory etiquette and hand hygiene. As it can be seen, social distancing but short of lockdown or stay at home orders. The evidence supporting the applications of such NPIs is the studies of the effect of NPIs on the mortality outcomes of different US cities during the outbreak of pandemic influenza (1918-1920).[14,15] Again, the NPIs were short of lockdown or stay at home order. Moreover, it is explicitly stated,[15] that an experimental study on the effect (and collateral effect) of NPI has not been performed because*:”the trend away from such traditional public health measures for disease control during the past 50 years*,(stated in 2007) *and ethical limitations of using population-wide nonpharmaceutical interventions in the absence of a serious threat*”.[15] It seems that COVID-19 is such a big threat that even harsher NPIs are considered adequate today.

A key factor is to be able to predict the importance of the threat, related with the number of cases or deaths.

Plenty of models have been set-up to model the number of cases and deaths of COVID-19. Most rely on SIR (Susceptible-Infected-Removed) simulation using systems of differential equations.[16] A more complex kind of models involves the so called agents, where the movements, transmission and infection of individuals are randomly produced and the infection rate calculated. Both kinds of models requires to know several parameters (e.g. number of persons infected by one infected individual) which can be estimated from earlier cases (e.g. Wuhan in China) but likely change from case to case, as it is shown in Europe. A well-known calibrated SRI simulation was made by the Imperial College Group,[16] whom inform policy in the UK, US and the world (including southern hemisphere countries (SHC)). They predict a maximum of deaths of 2.6M and 200K for USA and UK, respectively if “suppression” was not enacted. Mitigation measures which include isolation of cases and contacts, social distancing, ban of public gatherings and schools were considered insufficient.[18] Such dire forecast prompts both governments to apply social isolation. In that situation, the model predicts only 200K and 50K for the USA and UK, respectively. Most of other countries, including Argentina, followed suit and apply lockdown. In all cases was a quite informed decision because the ICG predicts 3.2M deaths for Latin America & Caribbean in the case no measures were taken, 729K if the “suppression” is made after 1.6 deaths/week/100K inhabitants has reached and 158K in case the “suppression” is enacted before 0.2 deaths/week/100 K has been reached. It should be mentioned that the last threshold means ca. 12 deaths per day in the whole Argentina. Such threshold have not been reached when total lockdown was enacted (23^rd^ of March 2020), was not reached since and the forecast presented here suggest that will never be reached.

On the other hand, the Institute for Health Metrics and Evaluation (IHME) of Washington University,[18] uses a fitting method to obtain a parametric equation which allows forecasting the number of deaths during time using the data of the ongoing outbreak. The equation that describes the number of deaths per day is the Gaussian distribution equation. The total number of deaths per day is the integral form of the equation. The equation represents a bell curve which is the shape found out empirically by Farr (Farr’s law) which have found to fit most of epidemic outbreaks of diseases.

While IMHE models several countries of the world, it does not apply the methodology to the southern hemisphere or the world, where Argentina is located. For informing health policy in our country, both our data and that of our neighbors is relevant.

Therefore, programs were set to fit the integral of the Gaussian equation. The fitting and forecasting capabilities of the programs were tested with advanced cases (in the northern hemisphere) where can be compared with IHME data. Then, were applied to southern hemisphere countries and also to the whole world.

The forecast for countries in the southern hemisphere show relatively low excess deaths. The calculation of different scenarios for infection rates also shows a relatively low infection rates. Possible causes are discussed.

## Experimental

The program was developed in Python 3.76 (https://www.google.com/search?client=firefox-b-d&q=python) in the IDE Spyder (https://www.spyder-ide.org/) launched by Anaconda (https://www.google.com/search?client=firefox-b-d&q=anaconda+python) on a Dell Latitude 3460 laptop (i3, 4 Gb of memory and 600 Gb of disk). The Gaussian equation (Farr’s law) was symbolically integrated with Euler (http://euler.rene-grothmann.de/index.html) by adding integration constant.

Method: fit with routines in Python (scipy.optimize, https://www.scipy.org/) in our own Python programs (available at **https://github.com/cesarbarbero/programas-para-predecir-COVID-19/**). The test suite was from Italy that contains more than 2/3 of the peak (deaths per day)-We also fit China that contains the whole forward peak. However, it does not contain the full data before the peak since the first reports are 17 deaths (in Wuhan). Moreover, recently (17/04/2020), they have incorporated 50% more deaths without information on the day of death. Therefore, the dataset of China is not usable.

The program predicts the same parameters using total data from Italy (53 days) or 35 days (non zero deaths for most SHC), showing that it is able to adequately predict the evolution of deaths over time.

Initially, the data from Argentina was obtained from the Ministry of Health of the Nation (MSN) by manual recording from daily reports (https://www.argentina.gob.ar/coronavirus/informe-diario).

Then, the data from most southern hemisphere countries were obtained from worldometer (https://www.worldometers.info/coronavirus/#countries) by reading the graphs in the webpage (see acknowledgements for the help in data collection).

However, the final runs were made with the data produced by John Hopkins University and stored in a github repository (https://github.com/CSSEGISandData/COVID-19/blob/master/csse_covid_19_data/csse_covid_19_time_series/time_series_covid19_deaths_global.csv#L17). In that way, the database is public and its quality is independently verified.

A special program: “Graphical fit” was used to obtain roughly correct parameters (a,b,c,norm) to be used as seed and as central points for the constraint of the curve fitting routine. By trial and error, guided by the understading of each parameter on the shape of the curve, the simulated data is graphed along the experimental data. When a reasonable fit is obtained, the parameters are copied in the curve fitting program as seed. The constraints of the curve fit are set around those values. The curve fitting program is run until the parameters are different to the constraints. The numerical and graphical output is directly written in the manuscript, without any data manipulation. The results of the fit (numerical data and plots) for each country are provided in the supplementary information and in github.

## Results and discussion

### Test the parametric function fitting with data from Italy and Spain

To test the ability of the program to forecast the evolution we used the dataset from Italy which has already cover ca. 2/3 of the peak.

In Figure 1 it is shown the sum of deaths during time from the 22^nd^ of January (first reported death in the world). Along it, it is shown the calculated fitted points, which closely follow the actual data. Using the parametric integrated Gaussian equation (PIGE), it is possible to forecast the evolution of the deaths. I The equation is then extrapolated to the whole width of the curve, showing that the number of deaths flatten out at large times.

**Figure 1.**
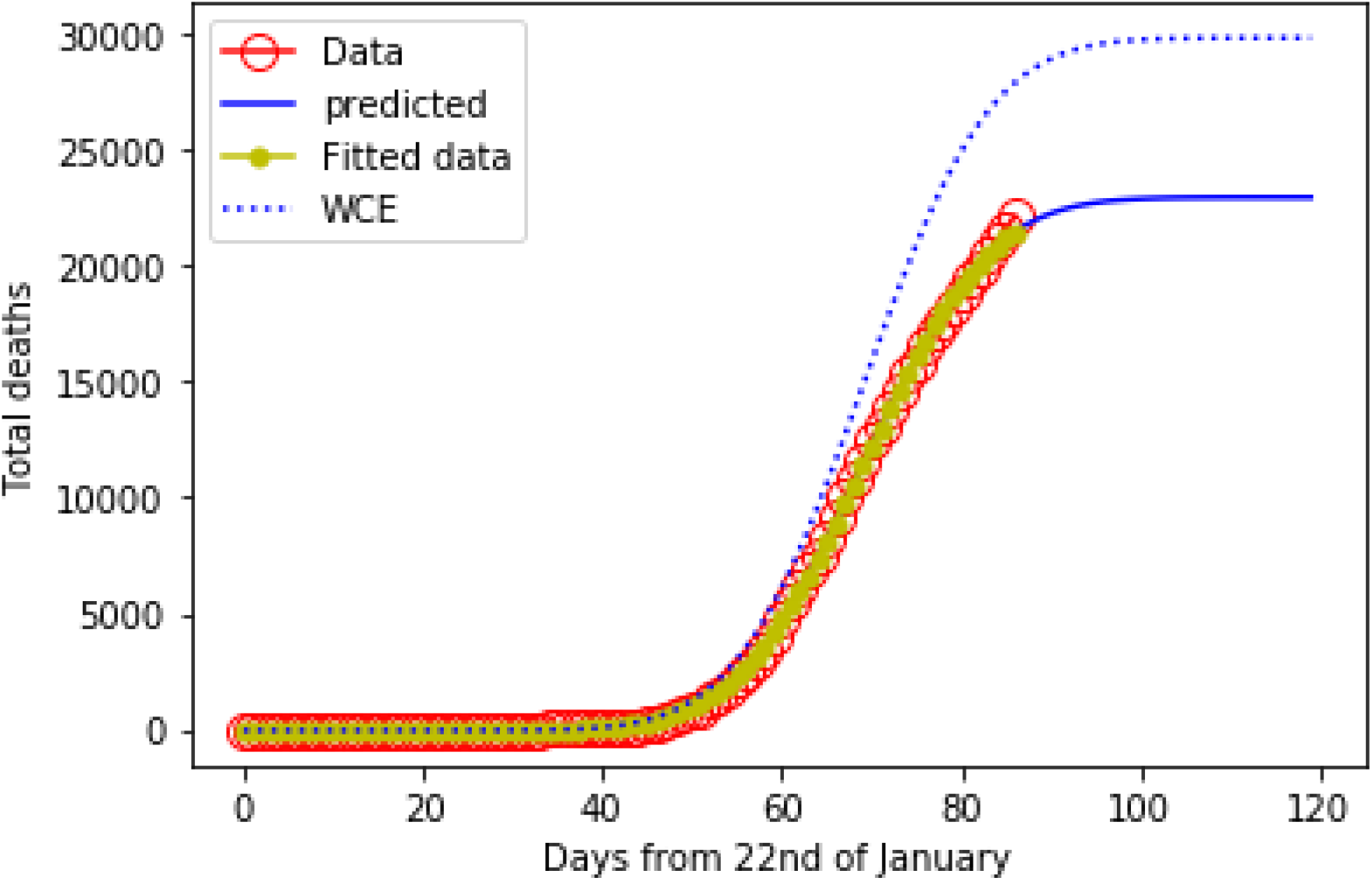
Plot of the total death by COVID-19 in Italy as a function of days elapsed the 22^nd^ of January(red circles). The yellow squares represent the fitted data while the blue line shows the predicted deaths by the parametric integrated Gaussian equation (PIGE) with the parameters produced by the fit. The dashed blue line represents the worst case scenario (WCE) with 50% uncertainty. In that way a total number of deaths of **22978** (29871-22978). The estimation of uncertainty is up to the 30% of the predicted data. Accordingly, in Figure 1 it is shown the worst case scenario (WCE) which correspond to the upper limit of the error. It should be noticed that the use of this uncertainty region in the graph of deaths per day allow to include the 95% of the scattered data (see below). The best case scenario (BCE) is not plotted in Figure 1 since it will show an unreal case of lower than actual deaths before the last real data point. It is likely for the deaths to be under-reported,^*^ but it is unlikely to be over-reported. Accordingly, the BCE for the total deaths (16804) is not shown because it is lower than the deaths already reported (22170 al 17-04-2020).The projected number of deaths agree (taken into account the uncertainty range) with the total number of deaths projected by the IHME of 25007 (31056-23589).[https://covid19.healthdata.org/italy accessed 18/04/2020)].

Using the parameters of the PIGE, it is possible to calculate the evolution of the number of deaths per day (parametric Gaussian equation, PGE) and compare with the reported data. In Figure 2 it is shown the graph of the number of deaths reported for Italy for each day. While the total number of deaths at each given day shows a smooth curve (Figure 1), the data of number of deaths per day show a large scattering. This is reasonable since the local reporting reach the health authorities at different rates, depending on distance of local governance. However, the predicted curve follow well the data and the uncertainty range (shown by dashed blue lines) of the forecast includes more than 95% of the reported data. In this case, the low range includes actual reported data.

**Figure 2.**
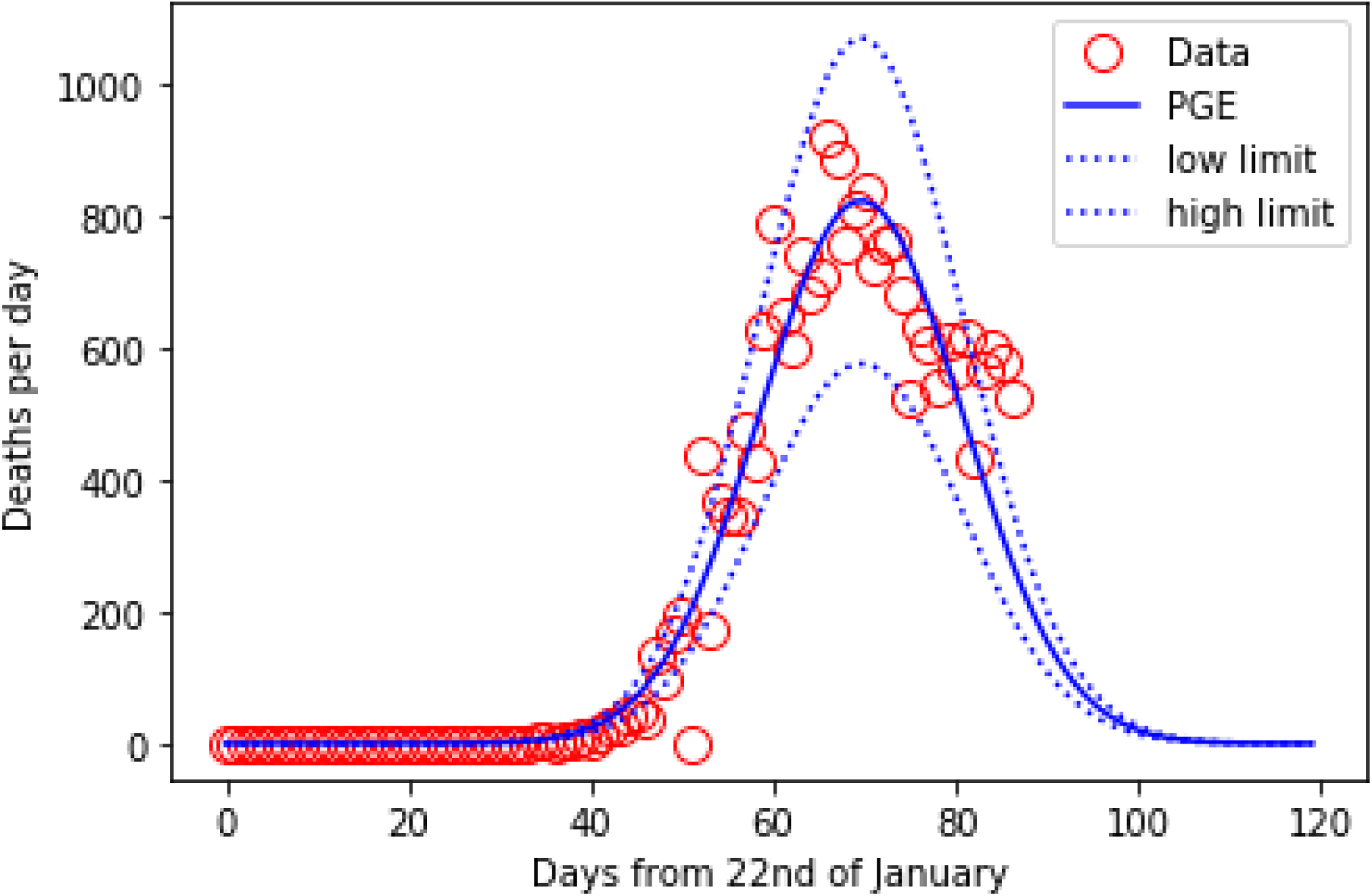
Graph of the deaths per day (red circles) along with the parametric Gaussian equation (PGE) plot calculated using the parameters obtained by the fit (blue line). The dashed curves showing the upper and lower limit of the uncertainty range (30% of PGE).

From the PGE it is possible to obtain other parameters of interest. First, it can be predicted the maximum of the peak, when the outbreak is at maximum which was de 1^st^ (+/-3) of April. As it can be seen in Figure 2 the reported deaths data show a clear maxima in that region.

More interesting is to predict the end of the outbreak. 100% of deaths will occur before the 16^th^ (+/-3) of May 2020. Using the 3 σ criterion (99.748 %), the outbreak will be the finished by 1^st^ (+/-3) of May. Note that the curve in Italy has already passed the σ point (68.27%) which happened the 5^th^ (+/-3) of April. Therefore, it is a convenient test suite for the fitting program.

The good fitting with Gaussian equation allows calculating a parameter which is relevant and has acquired growing epidemiological importance that is the true start of the outbreak. While in well-known diseases (e.g. measles) the cause of death is clearly assigned, with COVID-19 requires testing for the virus. In China the initial reports (22^nd^ of January) are of 17 cases. Applying the program to China show that there should be deaths due to COVID-19 before but there were not tests available. In other countries where the outbreak began later, the situation should be better. However, if the outbreak was not detected, deaths by COVID-19 could be assigned to other pathologies. Since the period between contagion an deaths is ca. 6 days [<u>Updated understanding of the outbreak of 2019 novel coronavirus (2019nCoV) in Wuhan, China - Journal of Medical Virology, Jan. 29, 2020</u>], it allow to find out the true beginning of the outbreak. In the case of Italy, the program calculates the 28^th^ (+/-3) of December 2019 as the date when 0% deaths has happened. Therefore, contagion in Italy should have been occurring in December 2019.

To test the capacity of the program to predict the evolution, the test was performed using data well before the peak (only till 21^st^ of March). The forecast shows similar outcomes predicting a total number of deaths of 25456 (33092 – 254569). The times of different events is even closer, with peak of death the 1^st^ (+/-3) of April, 100 % deaths the 20^th^ of May, σ day the 4 of April and beginning of the outbreak the 24^th^ of December 2019. Another test case used was Spain. The relevant parameters are depicted in Table 1. The detailed data with plots, similar to Fig. 1 and Dig. 2, are presented in the supplementary information.

**Table 1.** Relevant parameters of different countries in the Northern Hemisphere which has already overcome the peak of daily deaths, obtained by fitting the sums of daily death data.

| Country | Calculated deaths |  | Maximun<br>deaths/day | Dates<sup>§</sup> |  |  | Infected |  |
| --- | --- | --- | --- | --- | --- | --- | --- | --- |
|  | Max. | predicted |  | Begin | Peak | End | Max. | Min. |
| Italy | 29871 | 22978 | 823(+/-100) | 28-12-19 | 01-04-20 | 16-05-20 | 4595659 | 287228 |
|  | (26001)<sup>&</sup> |  |  |  |  |  |  |  |
| Spain | 26132 | 20102 | 902(+/-100) | 17-12-19 | 04-04-20 | 11-05-20 | 4020425 | 251276 |
|  | (23680)<sup>&</sup> |  |  |  |  |  |  |  |

<sup>§</sup> All dates are in the format dd/mm/yy.
<sup>&</sup> Total number of deaths predicted by IHME (https://covid19.healthdata.org/)

### Assessing relevant parameters of Argentina

The most personally relevant case for this study is Argentina. Moreover, the 23^rd^ of March was applied a total lockdown which is strongly affecting the economy and life in the country. Forecasting the magnitude of the problem is critical to advise policy.

The fitting of the reported total deaths data is shown in Figure 3.

**Figure 3.**
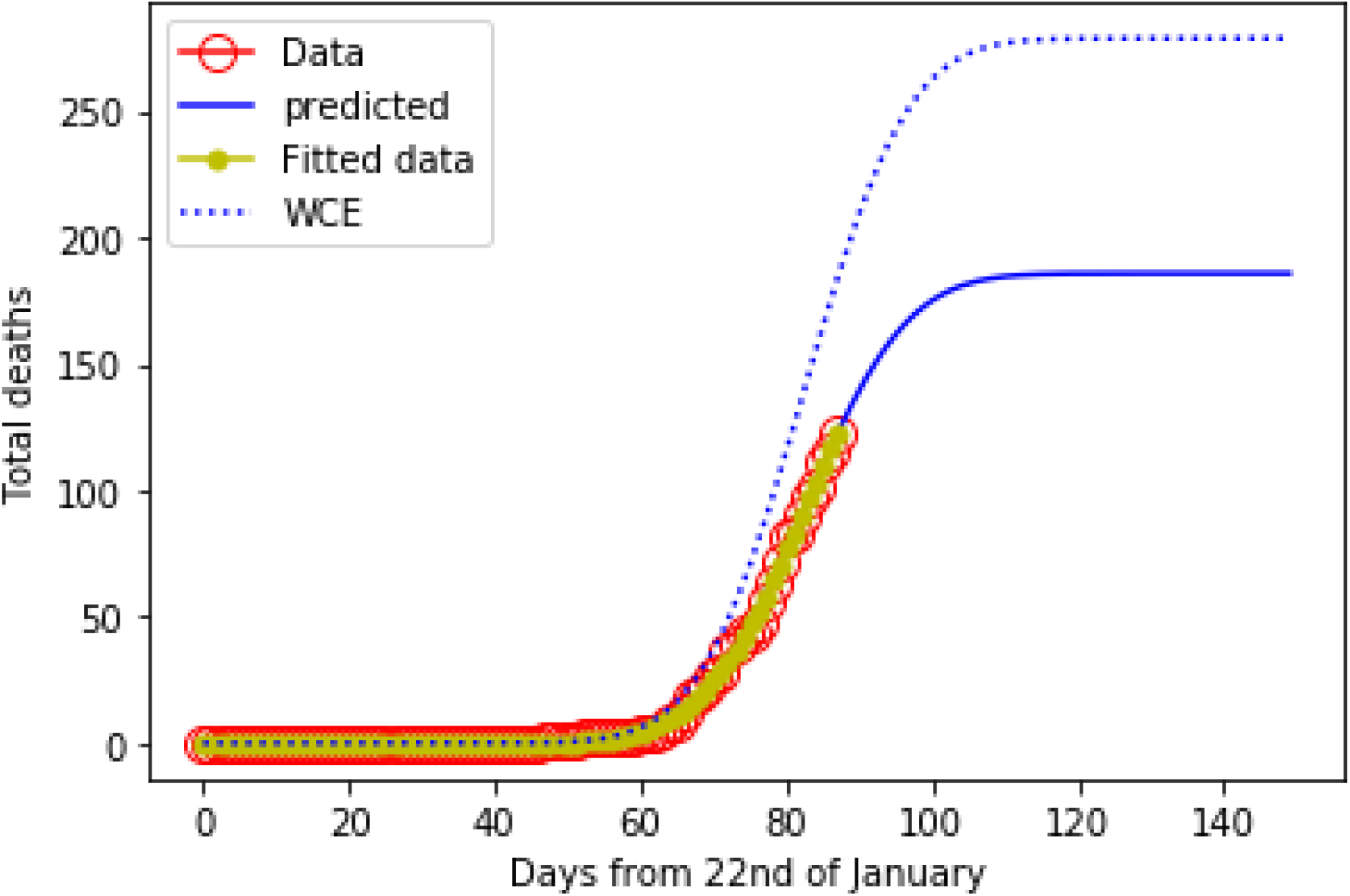
Plot of the total death by COVID-19 in Argentina as a function of days. The yellow squares represent the fitted data while the blue line shows the integral of the gaussian equation with the parameters produced by the fit. The dashed blue line plots the worst case scenario (with and added 50%) of death due to COVID-19 in Argentina.

**Figure 4.**
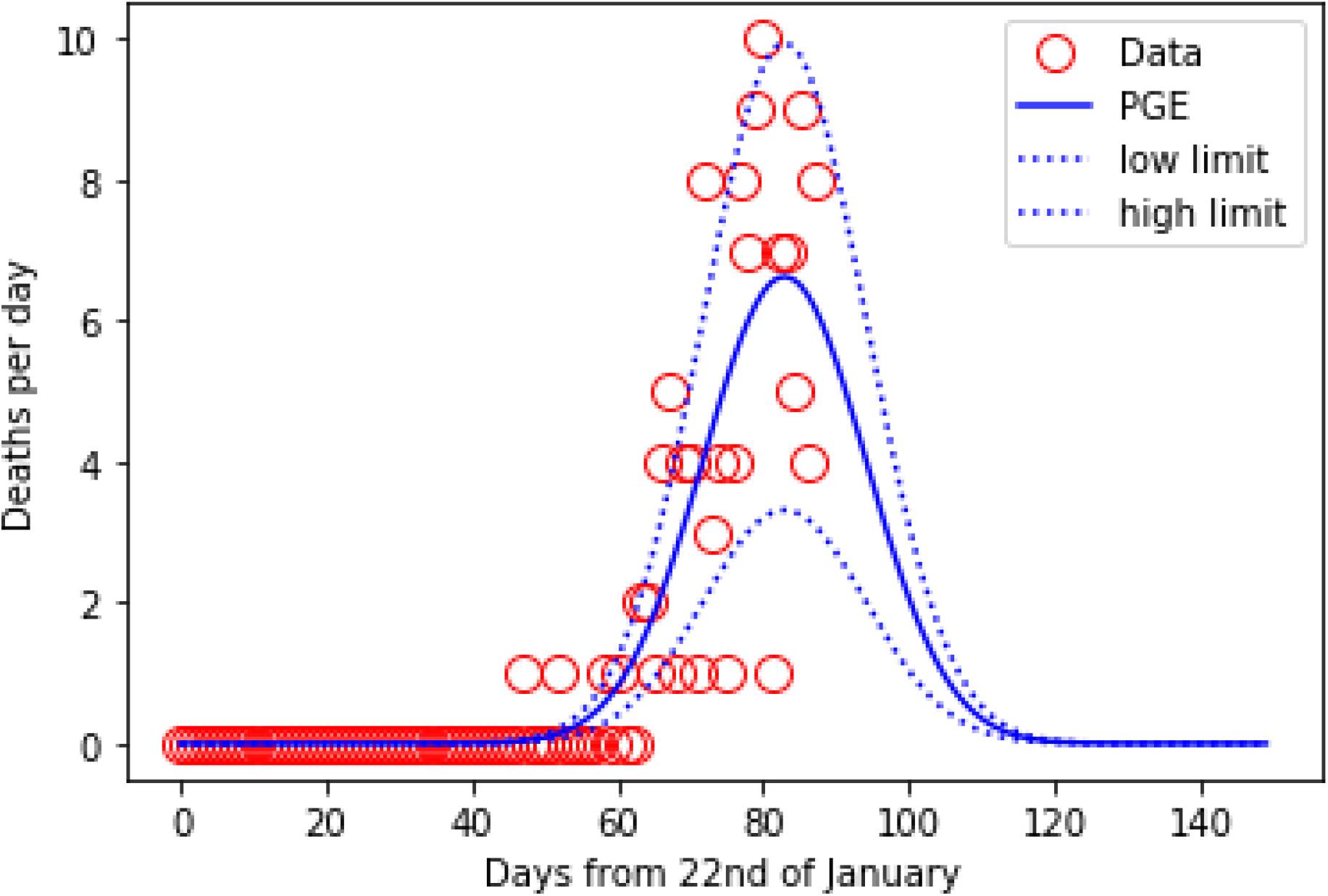
Graph of the deaths per day (red circles) along with the parametric gaussian equation plot calculated using the parameters obtained from the fit (blue line). The high and low limit of the uncertainty (30%) are shown as dashed blue lines.

The maximum number of deaths is found to be 182 (277-182) deaths, with a 50% of maximum uncertainty. The number is quite low, representing an excess death of 0,008 %. It is also lower than the number of people who died in 2009-2010 during the H1N1 influenza pandemic[18].

Other important parameters are the day of maximum death since it is related to the day of maximum usage of the health system. To find out, the Gaussian equation is plotted along with the data of deaths per day. (Figure 2).

In the case of Argentina, the maximum, 6 (+/-2), deaths happened the 14 (+/-3) of April. Note that the reported data of deaths per day show a large scatter (worse than Italy) due to the difficult reporting of few deaths in a large country. The large scatter makes difficult to fit directly the Gaussian equation but 95% of the actual point occurs inside the uncertainty range (50%).

Additionally, the equation allows calculating the end of the outbreak which is the 20^th^ (+/-3) of May. However, already the 1^st^ (+/-3) of May, 95.45% (2 σ) of the deaths have occurred. It is reasonable to think that at that date, the outbreak is over. Moreover, the parametrc equation allows to calculate the beginning of mortality due to COVID-9 which is the 9^th^ (+/-3) of March 2020.

As it can be seen, the PIGE predicts a relatively low number of deaths. The number for the Southern Hemisphere (last but one row at the left in Table 2) is significantly lower (10 to 15 times) than the projected best scenario (for Latin America & Caribbean only) in the ICG modeling (shown in the last row at the left of Table 2). The suppression imply 75% lockdown before the death rate has reached 0.2 deths/per week/100.000 inhabitants. [17].

**Table 2.** Relevant parameters of different countries in the Southern Hemisphere (SH) obtained by fitting the sums of daily death data.^†^ The countries are ordered in descending order, taking into account the projected total deaths. The dates are in the format dd/mm/yy.

| Country | Total deaths |  | Max. daily deaths | Calculated dates |  |  | Infected |  |
| --- | --- | --- | --- | --- | --- | --- | --- | --- |
|  | Max- | Projected |  | Begin | Peak | End | Max. | Min. |
| **Brazil** | 9157 | 6105 | 200 (+/-50) | 25-11-19 | 23-04-20 | 15-06-20 | 1221005 | 76312 |
| **Mexico** | 1860 | 1240 | 51(+/-10) | 18-03-20 | 21-04-20 | 25-5-20 | 248033 | 15502 |
| **Peru** | 712 | 475 | 19(+/-3) | 18-03-20 | 16-04-20 | 15-05-20 | 95069 | 5941 |
| **Colombia** | 300 | 450 | 12(+/-3) | 28-12-19 | 18-04-20 | 21-05-20 | 60018 | 3051 |
| **Ecuador** | 612 | 408 | 19(+/-3) | 08-12-19 | 08-04-20 | 20-04-20 | 81670 | 5104 |
| **South Africa** | 1113 | 371 | 17 | 20-03-20 | 14-05-20 | 08-07-20 | 148485 | 9280 |
| **Cuba** | 361 | 200 | 5(+/-2) | 02-04-20 | 10-05-20 | 17-06-20 | 48384 | 3024 |
| **Argentina** | 277 | 182 | 6(+/-2) | 09-02-20 | 14-04-20 | 20-05-20 | 36412 | 2275 |
| **Chile** | 231 | 154 | 7(+/-2) | 22-03-20 | 13-04-20 | 05-05-20 | 30937 | 1933 |
| **Panama** | 217 | 145 | 5 | 12-03-20 | 12-04-20 | 13-05-20 | 29039 | 1814 |
| **Australia** | 100 | 67 | 2 | 19-03-20 | 07-04-20 | 26-04-20 | 13593 | 849 |
| **Bolivia** | 54 | 36 | # | 08-03-20 | 14-06-20 | 25-04-20 | 7369 | 460 |
| **Honduras** | 52 | 35 | 1 | 15-03-20 | 07-04-20 | 30-04-20 | 7004 | 437 |
| **New Zealand** | 19 | 13 | 1 | 06-04-20 | 15-04-20 | 24-04-20 | 2606 | 162 |
| **Paraguay** | 18 | 12 | # | 19-03-20 | 13-04-20 | 08-05-20 | 2535 | 158 |
| **Guatemala** | 16 | 11 | # | 29-03-20 | 18-04-20 | 08-05-20 | 2346 | 146 |
| **Venezuela** | 13 | 9 | # | 24-03-20 | 03-04-20 | 13-04-20 | 1806 | 112 |
| **Uruguay** | 12 | 8 | # | 26-03-20 | 05-04-20 | 15-04-20 | 1605 | 100 |
| **El Salvador** | 9 | 6 | # | 31-03-20 | 06-04-20 | 12-04-20 | 1257 | 78 |
| **Jamaica** | 7 | 5 | # | 24-03-20 | 06-04-20 | 19-04-20 | 1111 | 69 |
| **Haiti** | 4 | 3 | # | 05-04-20 | 10-04-20 | 15-04-20 | 622 | 38 |
| **Costa Rica** | 4 | 3 | # | 21-03-20 | 31-03-20 | 10-04-20 | 774 | 48 |
| **Nicaragua** | \$ | n.d. | -- | -- | -- | -- | -- | -- |
| **SH=** | **15148** | **9939** |  |  |  |  | **2042419** | **126939** |
| **IGC\*=** | 158000 |  |  |  |  |  | 45346000 |  |

Not all the countries studied decide to apply a full lockdown (Chile for example apply a mitigation protocol in areas with few cases and good health coverage) and others have difficulties to enforce total lockdown (e.g. Brazil) but the outcomes are everywhere dramatically better than the IGC estimation. Therefore it is reasonable to assume that the IGC model overestimate the number of deaths.

The parametric equation allows detecting the maximum of deaths per day peak. Several countries have already passed the peak (including Argentina) meaning that the outbreak is subsiding. Since the number of deaths have to be related with the number of new infections (with a delay), it also means that number of new cases is decreasing. In any epidemiological model, this means that the number of infections caused by an infected individual is decreasing in time. This could be to the fact that the number of susceptible contacts is decreasing (because all are already infected or recovered) or that the number of contacts is decreasing with time. A way to calculate the number of persons infected involves dividing by the rate of mortality (Rm = deaths/infected). However, Rm is not easily known because it would require to know exactly the number of persons infected in all the countries involved, and such number depends on the testing capabilities and testing protocol. In Argentina, only individuals who show symptoms are tested, therefore asymptomatic infected individuals are not counted. Since different Rm values have been reported, the Rm reported by South Korea (0.5 %) was used to produce the high limit number and that of Italy (8 %) to calculate the lower limit. The results are shown in the last two columns of Table 2. The numbers for the whole SH are shown in the last but one row (right) of Table 2. In the worst case scenario ca. 2M individuals will be infected, which is quite low since the population of the SH is of more than 664M. Obviously, if the rate of mortality of COVID-19 is as low of the seasonal influenza (0.1%), the number of cases will be higher (ca. 10 M) but such scenario means that the COVID-19 is not a grave menace for general population, as the seasonal influenza, and the generalized lockdown is unjustified.

In the last row (right), it is shown the predicted number of cases by ICG.[17] The number is more than 45M, which is more than 20 times larger than the calculated here. Even if the rate of mortality is as low as that of seasonal influenza, the number of cases predicted by IGC is 4-5 times off. One of the more important concerns in this pandemic outbreak is the ability of different health systems to cope with the number of cases. Therefore, it is relevant to know the number of cases per day. In that way, it is possible to calculate the needs (hospital beds and intensive care unit (ICU) beds for each country (Table 3). To do that, we assume that 20% of the cases require hospitalization and 5% of the cases require ICU beds. Using the population data and following the ICG report,[17] it is possible to calculate the number of hospital beds (and ICU beds) available. To do that we use the worst case scenario that all SH countries are Low Middle Income Countries (LMIC), which gives 1.5 beds/1000 inhabitants. Then, the some category of LMIC gives 1.75 ICU beds/100 beds. The Lower Income Countries (e.g. Bolivia) in the list requires less than 10 beds and less than 2 ICU beds because has a mean of less than one deaths at the peak.

**Table 3.** Burden of the health system by COVID-19 and % of population affected.

| Country | Population | Beds | ICU beds | Beds | ICU beds | Deaths/100.000 |  | Infected (% pop) |  |
| --- | --- | --- | --- | --- | --- | --- | --- | --- | --- |
|  |  | required | required | available | available | Max | Min | Max | Min |
| Brazil | 212559417 | 8000 | 2000 | 106280 | 2126 | 4,31 | 2,87 | 0,574 | 0,036 |
| Mexico | 128932753 | 2040 | 510 | 64466 | 1289 | 1,44 | 0,96 | 0,192 | 0,012 |
| Peru | 32971854 | 760 | 190 | 16486 | 330 | 2,16 | 1,44 | 0,288 | 0,018 |
| Colombia | 50882891 | 480 | 120 | 25441 | 509 | 0,59 | 0,88 | 0,118 | 0,006 |
| Ecuador | 17643054 | 760 | 190 | 8822 | 176 | 3,47 | 2,31 | 0,463 | 0,029 |
| Cuba | 11326616 | 200 | 50 | 5663 | 113 | 3,19 | 1,77 | 0,427 | 0,027 |
| Argentina | 45195774 | 240 | 60 | 22598 | 452 | 0,61 | 0,40 | 0,082 | 0,005 |
| Chile | 19116201 | 280 | 70 | 9558 | 191 | 1,21 | 0,81 | 0,162 | 0,010 |
| Panama | 4314767 | 200 | 50 | 2157 | 43 | 5,03 | 3,36 | 0,673 | 0,042 |
| Australia | 25499884 | 80 | 20 | 12750 | 255 | 0,39 | 0,26 | 0,053 | 0,003 |
| Bolivia | 11673021 | <10 | <2 | 5837 | 117 | 0,46 | 0,31 | 0,063 | 0,004 |
| Honduras | 9904607 | 40 | 10 | 4952 | 99 | 0,53 | 0,35 | 0,071 | 0,004 |
| New Zealand | 4822233 | 40 | 10 | 2411 | 48 | 0,39 | 0,27 | 0,054 | 0,003 |
| Paraguay | 7132538 | <10 | <2 | 3566 | 71 | 0,25 | 0,17 | 0,036 | 0,002 |
| Guatemala | 17915568 | <10 | <2 | 8958 | 179 | 0,09 | 0,06 | 0,013 | 0,001 |
| Venezuela | 28435940 | <10 | <2 | 14218 | 284 | 0,05 | 0,03 | 0,006 | 0,000 |
| Uruguay | 3473730 | <10 | <2 | 1737 | 35 | 0,35 | 0,23 | 0,046 | 0,003 |
| El Salvador | 6486205 | <10 | <2 | 3243 | 65 | 0,14 | 0,09 | 0,019 | 0,001 |
| Jamaica | 2961167 | <10 | <2 | 1481 | 30 | 0,24 | 0,17 | 0,038 | 0,002 |
| Haiti | 11402528 | <10 | <2 | 5701 | 114 | 0,04 | 0,03 | 0,005 | 0,000 |
| Costa Rica | 5094118 | <10 | <2 | 2547 | 51 | 0,08 | 0,06 | 0,015 | 0,001 |
| Nicaragua | 6624554 | <10 | <2 | 3312 | 66 | .. | .. | ... | ... |
| **S.H. =** | **664369420** |  |  |  |  | **2,11** | **1,44** | **0,29** | **0,02** |

As it can be seen, only Peru, Ecuador and Panama should have not enough ICU beds to cope with grave COVID-19 patients.

The last four columns represent and evaluation of the proportion of population affected by COVID-19. The number of deaths per 100.000 individuals (per year) is below 5 while death by all causes is between 7000 and 11000. The percentage of population infected (calculated from mortality) will be less than 1%

## Conclusions

The fitting with a parametric integrated Gaussian equation (PIGE) seems to be a good method to obtain parameters of both descriptive and predictive value. Using open software libraries in Python (3.7) it is possible to produce programs which fit the experimental data (reported cumulative deaths). The programs are tested with advanced outbreaks (Italy and Spain) and give results comparable with those produced, using similar methods, by the IHME (Washington University, USA). The application to the reported data from Argentina allows forecasting a relatively small number of deaths (277-182). It also allows calculating: i) the day of the peak, the day of the begining and iii) the day of the end of the outbreak.

The application of the program to the data from the most important countries of the Southern Hemisphere shows a similar trend. While countries like Brazil and Mexico show more deaths it scales with a much larger population. In fact, relatively small countries (e.g. Uruguay) show so little number of deaths that makes the fitting difficult. The sum of deaths predicted for all the countries (even with an uncertainty of 50%) are significantly lower than those predicted by the Imperial College Group(ICG).

Using the parameterized death data and the mortality ratio (Rm), it is possible to calculate the number of COVID-19 cases, assuming two scenarios: a low Rm (South Korea) and a high Rm (Italy). These assumptions translate into a worst case scenario and best case scenario, respectively, for the number of infected individuals. These data are also much smaller than those predicted by the ICG.

Using the maximum deaths per day, calculated by the parameterized Gaussian equation (PGE) with the parameters obtained from the fit of PIGE, it is possible to forecast the maximum need of hospital and ICU beds. Using the assumptions of ICG, we can calculate the hospital and ICU beds available in each country. The comparison allows to state that all countries have enough hospital beds but Peru, Ecuador and Panama should have not enough ICU beds to cope with grave COVID-19 patients.

One possible reason of low mortality involves the fact that the outbreaks begun during late summer in the SH, instead or late winter as in the Northern Hemisphere. Therefore, a new harsher outbreak could occur after June (winter in the SH). If that is the case, the harsh lockdown measures implemented in late March would be a hard cure for a mild disease but it would made more difficult for SH countries to reapply such hard measures during winter when the new, potentially harder, outbreak develops.

## Data Availability

All datasets, programs and results are freele available from the author

https://github.com/cesarbarbero/programas-para-predecir-COVID-19/

## Acknowlegements

This work was funded by Universidad Nacional de Rio Cuarto (UNRC, Argentina) and the Consejo Nacional de Investigaciones Cientificas y Tecnicas (CONICET, Argentina) whom paid my salary while I am socially isolated at home. Special thanks to members of my research group: A. Cuello, D.Acevedo, E. Yslas, R. Bellingieri, R. Gramaglia, G. Morales, J. Balach and M.A. Molina whom helped with the initial data collection. Helpful virtual discussions with J. Balach and E. Quinteros about the program are also discussed. The laptop used was purchased with funds from a Secretaria de Politicas Universitarias – Minsterio de Educacion (Argentina). The author appreciates the use of open source programs (Python, Anaconda, Spyder, Matplotlib, Scipy, Numpy, Euler Math Toolbox, Vseuz graphics software). Finally, special thanks to Dr. L. Sereno whom teach me how to visually find out good seed/constraints for nonlinear fitting of complex equations.

## Supplementary information

A set of document files with the results of all the countries described and representative datasets. The Programs could not be loaded in medaRXiv. Are available at https://github.com/cesarbarbero/programas-para-predecir-COVID-19/).

## Footnotes

* The province of Hubei (PRC) reported 1290 unreported deaths the 17th of April, which represent ca. 50% of all cases. (https://www.worldometers.info/coronavirus/country/china/ accessed 17/04/2020.

† Downloaded from github the 18-04-2020, except Argentina which is updated on 20-04-2020

## Notes

### Competing Interest Statement

The authors have declared no competing interest.

### Funding Statement

UNRC-CONICET

